# Diagnosis in children with prolonged or recurrent cough: findings from the Swiss Paediatric Airway Cohort

**DOI:** 10.1101/2024.01.21.24301573

**Authors:** Maria Christina Mallet, Annina Elmiger, Sarah Glick, Tayisiya Krasnova, Carmen CM de Jong, Barbara Kern, Alexander Moeller, Nicolas Regamey, Oliver Sutter, Jakob Usemann, SPAC Study Team, Eva SL Pedersen, Claudia E Kuehni

**Affiliations:** Institute of Social and Preventive Medicine, University of Bern, Bern, Switzerland; Graduate School for Health Sciences, University of Bern, Bern, Switzerland; Division of Paediatric Respiratory Medicine, Department of Paediatrics, Inselspital, Bern University Hospital, University of Bern, Bern, Switzerland; Department of Paediatrics, Kantonsspital Aarau, Aarau, Switzerland; Department of Respiratory Medicine, University Children’s Hospital Zurich and Children’s Research Center, University of Zurich, Zurich, Switzerland; Division of Paediatric Pulmonology, Children’s Hospital, Cantonal Hospital Lucerne, Lucerne, Switzerland; Paediatric Practice, Worb, Switzerland; University Children’s Hospital Basel UKBB, Basel, Switzerland; A list of the SPAC Study Team can be found in the acknowledgements section

**Keywords:** Children, chronic cough, prolonged cough, recurrent cough, diagnosis, respiratory outpatient clinics

## Abstract

**Introduction:** Prolonged or recurrent cough is a common reason for referral to pediatric pulmonologists, yet few studies have assessed its causes. We examined records of children visiting respiratory outpatient clinics in Switzerland and assessed how diagnoses vary by age.

**Methods:** We analyzed data from the multicenter Swiss Paediatric Airway Cohort study. We included 363 children (median age 6 years, range 0–16) referred for prolonged or recurrent cough. From outpatient records, we extracted information on diagnostic investigations, final diagnoses proposed by pediatric pulmonologists, and treatments prescribed.

**Results:** Asthma and asthma-like conditions (cough variant asthma, episodic viral wheeze, and recurrent obstructive bronchitis) was diagnosed in 132 (36%) of 363 children, respiratory tract infections (RTI) including protracted bacterial bronchitis (PBB) in 51 (14%), upper airway cough syndrome (UACS) in 48 (13%), postinfectious cough in 36 (10%); other diagnoses including gastroesophageal reflux disease (GERD) and somatic cough syndrome or tic cough were found in 23 (6%). No etiology was found in 73 children (20%). Asthma was diagnosed 3.5 times more often in schoolchildren while RTI including PBB was diagnosed 3 times more often in preschoolers. Inhaled corticosteroids were prescribed for 84% of children diagnosed with asthma and asthma-like conditions, antibiotics for 43% of children with RTI, and nasal corticosteroids for 83% of those with UACS.

**Conclusion:** Coughing children received a wide spectrum of diagnoses that differed between preschool and schoolchildren. Asthma accounted for 36% of diagnoses, which emphasizes the importance of comprehensive investigation beyond asthma in children with prolonged or recurrent cough.

## 1. INTRODUCTION

Prolonged or recurrent cough is a common presenting symptom in pediatric primary care and a frequent reason for referral to pediatric pulmonologists [1-3]. The spectrum of differential diagnoses is broad and ranges from asthma and asthma-like conditions (i.e. cough variant asthma, episodic viral wheeze, and recurrent obstructive bronchitis), postinfectious cough, protracted bacterial bronchitis (PBB), upper airway cough syndrome (UACS), gastroesophageal reflux disease (GERD), somatic cough syndrome (previously psychogenic cough), and tic cough (previously habit cough) to rarer and more serious respiratory conditions such as cystic fibrosis (CF), primary ciliary dyskinesia (PCD), or primary immunodeficiency [1, 4, 5]. Establishing the underlying etiology of prolonged or recurrent cough can be challenging, and even after thorough investigation a cause is not always found [6]. Yet identifying the cause of prolonged or recurrent cough, making a correct diagnosis, and prescribing the adequate treatment is important because cough affects the quality of life of children and their families and burdens the healthcare system [2, 7, 8]. A systematic review of clinical studies reported that the most frequent causes of chronic cough in children were asthma and PBB, but the frequency of causes varies with age and across healthcare settings [9], and in population-based studies asthma as a frequent cause of cough was found to be inconsistent [10-12]. Causes of cough in children also differ from those in adults. In young children, common causes include postinfectious cough, PBB, and asthma, while in older children causes include asthma, UACS, and GERD and are similar to those in adults [7, 13].

Studies assessing causes of chronic cough in children visiting healthcare settings are limited [14]. Most research had been conducted in the United States, Australia, Turkey, and China [1, 15-27]. Many studies had small sample sizes (< 100 patients) [3, 15-17, 25, 27], included only patients from highly specialized centers [15, 17], did not assess how etiologies differ by age or only included children within a restricted age range [3, 16-21, 25-27], and most were single-center studies with limited generalizability [3, 15-27].

Drawing upon data from children referred to respiratory outpatient clinics in Switzerland for evaluation of prolonged or recurrent cough, we examined diagnostic investigations, final diagnoses, and the treatments prescribed in clinical settings. Based on previous clinical studies [1, 3, 18-23, 26], we hypothesized that asthma and PBB would be the most common diagnoses and that the relative frequency of diagnoses would vary by age.

## 2. METHODS

### 2.1 Study design and population

The Swiss Paediatric Airway Cohort (SPAC) is a prospective, national, multicenter clinical cohort of children in Switzerland. The design and methods of SPAC have been described in detail elsewhere [28]. SPAC enrolls children aged 0–16 years who visit respiratory outpatient clinics or pediatric pulmonary practices after being referred by general practitioners or pediatricians for the diagnostic evaluation of common respiratory problems such as cough, wheeze, and exercise-induced symptoms. Children with a prior diagnosis of a severe lung disease such as CF or PCD, cardiac problems, or neuromuscular disorders are excluded from the study. SPAC was set up in 2017 and recruitment is ongoing in eight pediatric respiratory outpatient clinics and two pediatric pulmonary practices. The study is observational and integrated into the routine care of the participating centers; therefore, no diagnostic evaluations are performed specifically for the SPAC study, and objective diagnostic tests and treatments are based on local protocols and indications. The Bern Cantonal Ethics Committee (Kantonale Ethikkommission Bern 2016–02176) approved the study; written informed consent was obtained from parents and patients age ≥ 14 years.

### 2.2 SPAC study procedures and data sources

During their visit, eligible patients are invited to enroll and parents complete a baseline questionnaire with information on respiratory symptoms, comorbidities, personal and family history, environmental exposures, and sociodemographic information. After the visit, the SPAC study team collects information from the patients’ medical records that includes referral letters with information about reasons for referral and outpatient clinical letters with information about diagnostic investigations performed, diagnosis, and treatment prescribed.

### 2.3 Inclusion criteria

For this analysis, we considered children who were invited to participate in the SPAC study from July 2017 until September 2022. We screened participants through referral and initial outpatient clinical letters of patients who had a signed informed consent and a completed baseline questionnaire. We included children newly referred for prolonged or recurrent cough as the main or only reason stated in the referral letter. We did not restrict our inclusion criteria to cases where the specific term "chronic cough"’—that is, a cough by definition lasting more than four weeks [4, 7, 29]—was used. This is because cough was heterogeneously described in the referral and outpatient clinical letters, and we wanted to capture all children presenting with any troublesome and persistent cough to a specialist consultation. Hence, we included all the children in whom the cough was described as “recurrent,” “recurrent prolonged,” “chronic recurrent,” “prolonged,” “chronic,” “protracted,” “persistent,” or “long-standing.” We refer to prolonged or recurrent cough throughout for simplicity. In the absence of a referral letter, we used the reason for referral given in the outpatient clinical letter. We excluded children who already had a confirmed asthma diagnosis before referral to the outpatient clinic.

### 2.4 Diagnosis given at the clinic

We collected information about the primary suspected or confirmed diagnosis for cough from the outpatient letter written by the attending pediatric pulmonologist. Because some children needed more than one visit for the diagnostic evaluation, we collected diagnoses from the most recent outpatient letter. We grouped diagnoses into five groups for the analysis: (i) asthma and asthma-like conditions (i.e. cough variant asthma, episodic viral wheeze, and recurrent obstructive bronchitis); (ii) nonspecific postinfectious cough with hyperactivity of cough receptors and/or bronchial hyper-reactivity, which we refer to simply as postinfectious cough; (iii) respiratory tract infections (RTI) including recurrent RTI and PBB; (iv) UACS previously known as postnasal drip; (v) unknown/unclear etiology and other diagnoses including GERD, somatic or tic cough, and other rarer diagnoses.

### 2.5 Parent-reported characteristics, diagnostic investigations, and treatment prescribed

We compared the diagnostic groups with respect to the information reported by parents in the baseline questionnaire: sociodemographic factors (sex, nationality, parental education), exposure to tobacco smoke, parental history of atopic diseases and cough, cough characteristics, and other respiratory symptoms in the past 12 months. We extracted information on diagnostic investigations from the outpatient letters. Diagnostic investigations were performed according to recommended guidelines [30-32] and included spirometry, body plethysmography, bronchodilator response test, bronchial challenge tests such as methacholine provocation and exercise challenge test, fractional exhaled nitric oxide (FeNO), nasal nitric oxide (nNO), allergy tests (skin prick test or specific IgE), radiological examinations (chest X-ray or computed-tomography [CT] scan), microbiology tests, and sweat test. Prescribed treatment was also extracted from the outpatient letters. We summarized treatment recommendations as asthma treatment including bronchodilators (e.g., short acting beta agonists [SABA]) and inhaled corticosteroids (ICS) alone and in combination with long-acting bronchodilators (LABA), leukotriene receptor antagonist (LTRA), oral corticosteroids, antihistamines, antibiotics, antireflux medications, nasal corticosteroids, and antitussives.

### 2.6 Statistical analysis

We report diagnoses, parent-reported information, diagnostic investigations, and treatment prescribed using proportions and percentages for categorical variables, and median and interquartile range (IQR) for continuous variables. We compared characteristics, diagnostic tests, and treatments by diagnostic groups using chi-squared or Fisher’s exact tests. We stratified the analyses for diagnosis given and diagnostic tests performed by age—preschool (< 5 years) and school-age (≥ 5 years)—since the maturity of the immune system and ability to correctly perform certain tests (e.g., lung function tests) differ.

Symptoms reported by parents in the baseline questionnaire had few missing values, ranging from 0 to 6%, except for cough triggers, 9 to 25% of which were missing (Table S1). We recoded missing values for questions on symptoms as "no" under the assumption that parents would have answered "yes" if symptoms were present and severe. As sensitivity analysis, we did a complete case analysis. We used STATA (Version 15.1, StataCorp) for the analysis.

## 3. RESULTS

Among the 2206 children with informed consent, a completed baseline questionnaire, and an initial outpatient clinical letter available, 363 (16%) had prolonged or recurrent cough as the main reason for referral (Figure S1). Among those, 218 (60%) were male and their median age was 6 years (IQR 4–8).

### 3.1 Diagnoses

The final diagnoses given by pediatric pulmonologists for children referred for prolonged or recurrent cough were diverse, heterogeneously described, and differed between preschool and school-age children (p < 0.001) (Figure 1 and Table S2 for details). The most frequent diagnoses among the study’s 363 children were asthma and asthma-like conditions in 132 (36%), followed by RTI in 51 (14%), which included recurrent RTI in 25 (7%) and PBB in 26 (7%), UACS in 48 (13%), and postinfectious cough in 36 (10%). Other diagnoses such as gastroesophageal reflux or somatic/tic cough occurred in only 23 children (6%). In 73 children (20%), the etiology and hence the final diagnosis remained unclear at the time of data collection.

**Figure 1:**
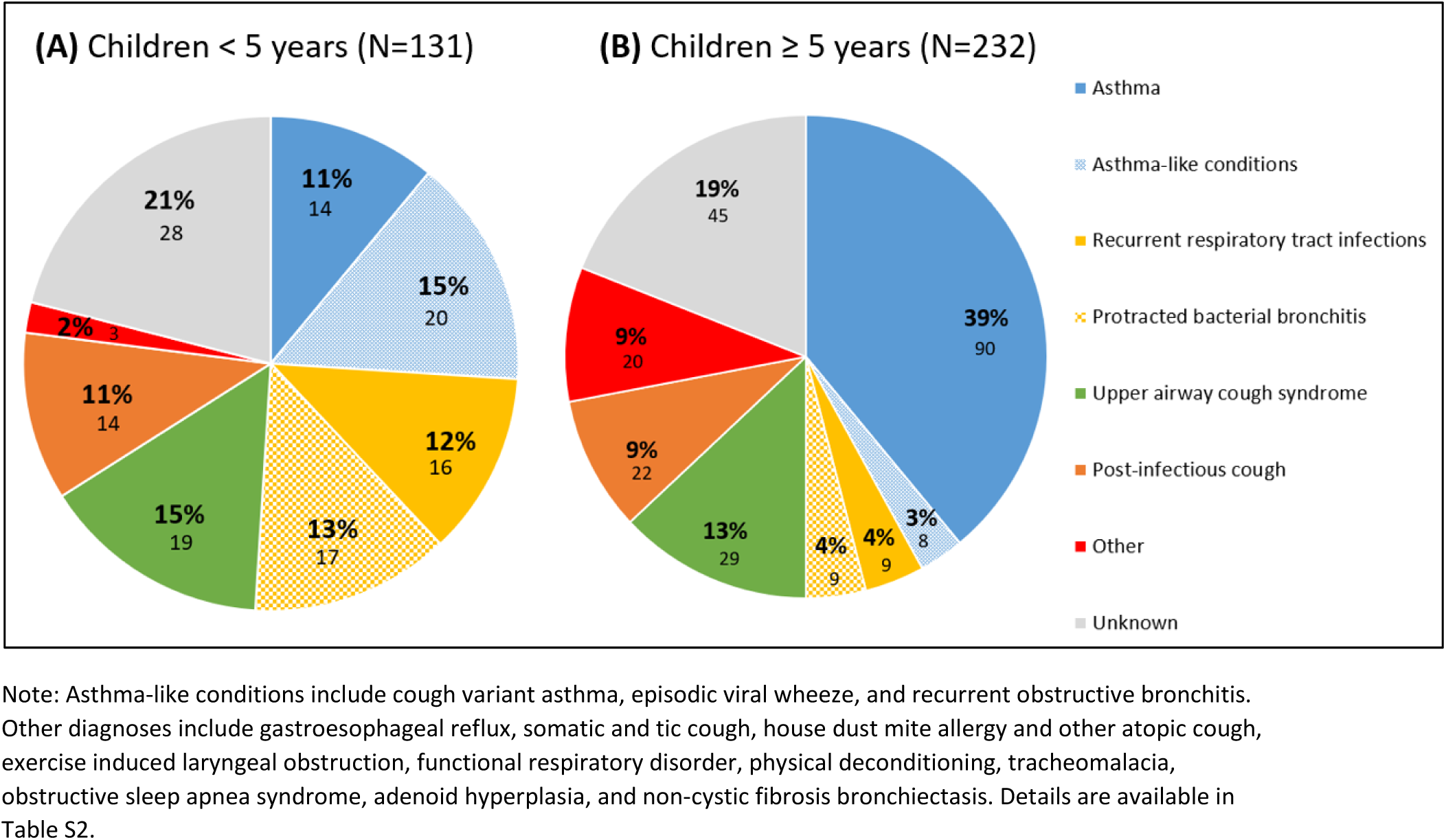
Percentage and number of final primary diagnosis given by pediatric pulmonologists to children seen for prolonged or recurrent cough by age group: (A) 131 children < 5 years and (B) 232 children ≥ 5 years

Among the 232 school-age children, asthma was diagnosed in 90 (39%), UACS in 29 (13%), postinfectious cough in 22 (9%), RTI in 18 (8%), and asthma-like conditions in 8 (3%). Among the 131 preschool children, 33 (25%) had RTI, with PBB comprising half of those diagnoses and 20 (15%) were found to have asthma-like conditions, which were diagnosed 3 to 5 times more often in preschoolers than in schoolchildren, while asthma was diagnosed in 14 preschool children (11%), a frequency that is about one-third that of asthma diagnoses among school-age children.

### 3.2 Comparison of parent-reported characteristics and diagnosis

Sociodemographic characteristics (sex, nationality, and parental education), exposure to tobacco smoke, and parental history of atopic diseases and cough did not differ among the diagnostic groups (Table 1). Seventy-eight percent of the children experienced cough lasting more than four weeks. This proportion ranged between 69% and 83% across the different diagnoses. Almost two-thirds of the children diagnosed with postinfectious cough or unknown/other etiology had a cough lasting more than two months. Cough with colds, cough without a cold, and coughing more than others were reported in most children (> 75%) across all diagnoses. Dry night cough was least often reported in children with RTI (57%) compared to other groups (≥ 70%). Parents described cough as mostly wet or both dry and wet in 40 of the 51 children diagnosed with RTI (78%) while it was mostly dry in more than half of the 48 children diagnosed with UACS or the 96 children in the unknown/other group. Parents of 172 children (47%) reported that their child had wheezed in the past 12 months. This proportion was highest in children later diagnosed with asthma and asthma-like conditions (62%). Frequent colds (≥ 7/year) were more often reported in children diagnosed with RTI (51%), snoring in UACS (52%), and rhino-conjunctivitis in children with asthma and asthma-like conditions (31%). In the complete case sensitivity analysis, the results differed minimally, and the overall conclusion remained the same (Table S3).

**Table 1:** Parent-reported characteristics and symptoms in the entire study population and stratified by diagnostic group.

|  | Total | Asthma and<br>asthma-like<br>conditions | Recurrent/<br>protracted<br>infection | Upper<br>airway<br>cough<br>syndrome | Post-infectious<br>cough | Unknown<br>etiology or<br>other | p-value |
| --- | --- | --- | --- | --- | --- | --- | --- |
|  | (N=363) | (N=132) | (N=51) | (N=48) | (N=36) | (N=96) |  |
|  | N (%) | N (%) | N (%) | N (%) | N (%) | N (%) |  |
| **Sociodemographic characteristics** |  |  |  |  |  |  |  |
| Male sex | 218 (60) | 85 (64) | 29 (57) | 26 (54) | 18 (50) | 60 (63) | 0.448 |
| Age, years (median, IQR) | 6 (4 – 8) | 6 (5 – 9) | 4 (2 – 6) | 5 (3 – 8) | 6 (4 – 8) | 7 (4 – 9) | <0.001* |
| Swiss nationality | 284 (78) | 102 (77) | 42 (82) | 35 (73) | 28 (78) | 77 (80) | 0.809 |
| Parental highest education |  |  |  |  |  |  |  |
| Maternal tertiary (N=353) | 124 (35) | 35 (27) | 23 (45) | 17 (38) | 13 (37) | 36 (38) | 0.182 |
| Paternal tertiary (N=350) | 154 (44) | 51 (41) | 22 (43) | 22 (49) | 15 (42) | 44 (47) | 0.833 |
| **Environmental exposure** |  |  |  |  |  |  |  |
| Parental smoking (N=351) | 125 (36) | 44 (34) | 19 (38) | 12 (27) | 15 (44) | 35 (38) | 0.579 |
| **Parental history** |  |  |  |  |  |  |  |
| Asthma | 115 (32) | 45 (34) | 17 (33) | 14 (29) | 12 (33) | 27 (28) | 0.884 |
| Hay fever | 166 (46) | 60 (45) | 19 (37) | 23 (48) | 17 (47) | 47 (49) | 0.734 |
| Chronic cough | 55 (15) | 22 (17) | 8 (16) | 5 (10) | 7 (19) | 13 (14) | 0.774 |
| **Cough symptoms and characteristics in the past 12 months** |  |  |  |  |  |  |  |
| Cough with colds | 355 (98) | 128 (97) | 50 (98) | 46 (96) | 35 (97) | 96 (100) | 0.251# |
| Cough without a cold | 319 (88) | 114 (86) | 39 (76) | 45 (94) | 31 (86) | 90 (94) | 0.024 |
| Dry night cough | 262 (72) | 92 (70) | 29 (57) | 37 (77) | 29 (81) | 75 (78) | 0.042 |
| Cough more than others | 328 (90) | 119 (90) | 45 (88) | 47 (98) | 29 (81) | 88 (92) | 0.099# |
| Cough > 4 weeks | 282 (78) | 99 (75) | 35 (69) | 40 (83) | 30 (83) | 78 (81) | 0.266 |
| Cough > 2 months | 178 (49) | 42 (32) | 26 (51) | 29 (60) | 23 (64) | 58 (60) | <0.001 |
| Cough type (N=362) |  |  |  |  |  |  |  |
| Mostly dry | 157 (43) | 54 (41) | 11 (22) | 26 (54) | 15 (42) | 51 (53) | <0.001 |
| Mostly wet | 44 (12) | 11 (8) | 18 (35) | 4 (8) | 3 (8) | 8 (8) |  |
| Both dry and wet | 161 (44) | 66 (50) | 22 (43) | 18 (38) | 18 (50) | 37 (39) |  |
| Cough triggers |  |  |  |  |  |  |  |
| Any aeroallergen | 126 (35) | 63 (48) | 11 (22) | 15 (31) | 6 (17) | 31 (32) | 0.001 |
| Any physical trigger (exercise/laughter/crying) | 273 (75) | 101 (77) | 37 (73) | 36 (75) | 26 (72) | 73 (76) | 0.971 |
| Cold air/temperature change | 196 (54) | 74 (56) | 25 (49) | 28 (58) | 23 (64) | 46 (48) | 0.413 |
| **Wheeze & other symptoms in past 12 months** |  |  |  |  |  |  |  |
| Wheeze | 172 (47) | 82 (62) | 24 (47) | 20 (42) | 12 (33) | 34 (35) | <0.001 |
| Number of colds ≥7/year | 111 (30) | 30 (23) | 26 (51) | 17 (35) | 10 (28) | 27 (28) | 0.005 |
| Otitis media | 64 (18) | 21 (16) | 13 (25) | 11 (23) | 4 (11) | 15 (16) | 0.322 |
| Snoring° | 144 (40) | 56 (42) | 12 (24) | 25 (52) | 13 (36) | 38 (40) | 0.055 |
| Pneumonia | 44 (12) | 18 (14) | 6 (12) | 3 (6) | 4 (11) | 13 (14) | 0.725 |
| Rhino-conjunctivitis | 77 (21) | 41 (31) | 2 (4) | 8 (17) | 4 (11) | 22 (23) | 0.001 |

°Without cold/almost always; p-values calculated using chi-square test; \*p-value calculated with kwallis test; #p-value calculated with Fishers exact test

### 3.3 Diagnostic investigations

The diagnostic investigations performed in the clinics varied across diagnostic groups and between preschool and school-age children. Overall, the three tests most commonly done were FeNO (73%), lung function tests including spirometry and body plethysmography (71%), and allergy tests (70%). FeNO and lung function were performed 115 times in the 132 children later diagnosed with asthma or asthma-like conditions (87%, each) and 68 times in the 96 children with an unknown/other etiology (71%, each; Table 2). A chest X-ray was done in about one-third of the 183 children diagnosed with postinfectious cough, RTI, or unknown/other etiology. Three CT scans were done, each in a child with an unknown etiology. As expected, lung function and FeNO were measured in almost all school-age children, but rarely in preschoolers (Table 3). Chest X-rays were done about three times more often in preschool than in school-age children (43% vs 16%).

**Table 2:**
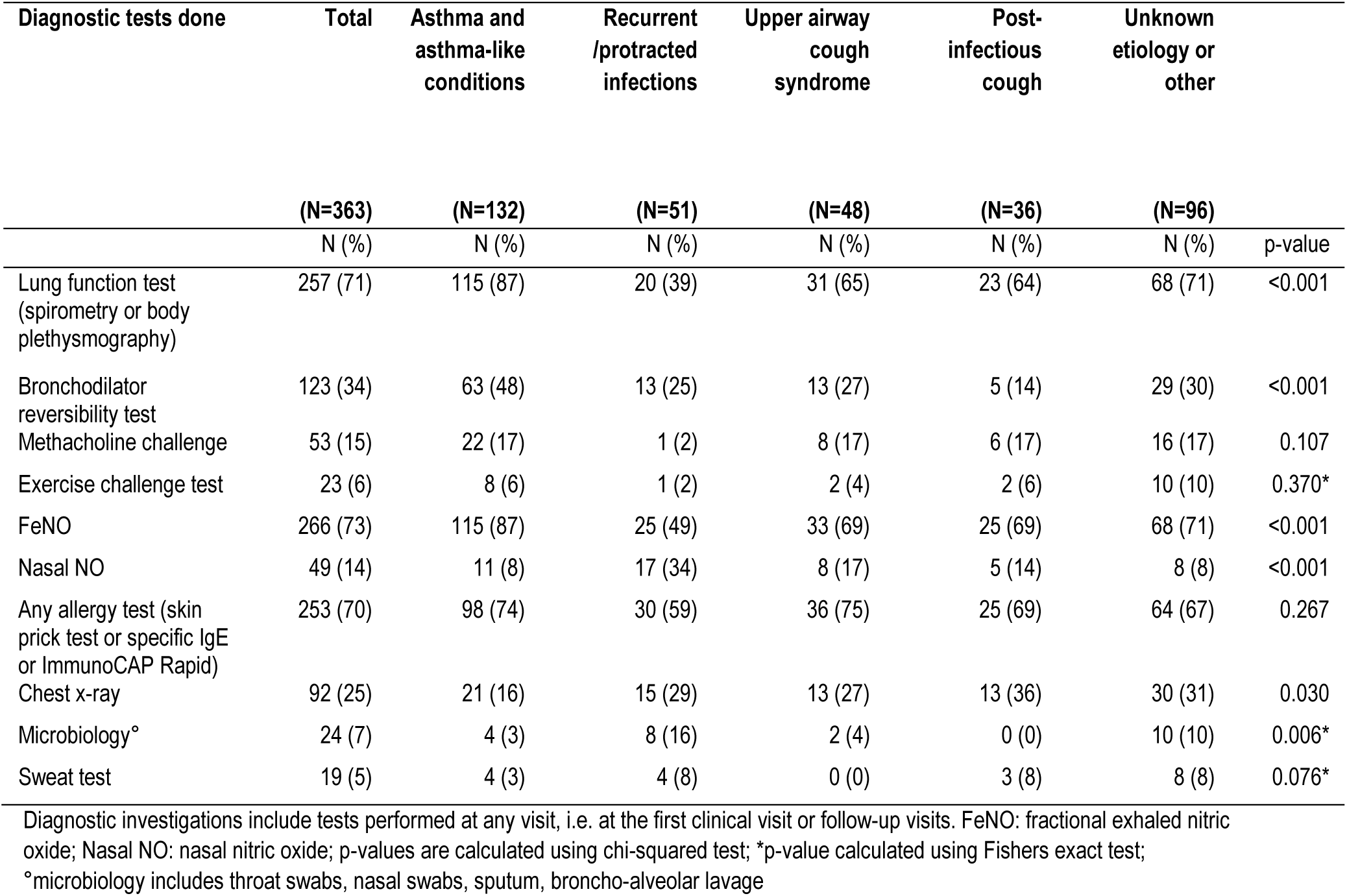
Diagnostic investigations performed in the entire study population and stratified by diagnostic group.

| Diagnostic tests done | Total | Asthma and asthma-like conditions | Recurrent /protracted infections | Upper airway cough syndrome | Post-infectious cough | Unknown etiology or other |  |
| --- | --- | --- | --- | --- | --- | --- | --- |
|  | (N=363) | (N=132) | (N=51) | (N=48) | (N=36) | (N=96) |  |
|  | N (%) | N (%) | N (%) | N (%) | N (%) | N (%) | p-value |
| Lung function test (spirometry or body plethysmography) | 257 (71) | 115 (87) | 20 (39) | 31 (65) | 23 (64) | 68 (71) | <0.001 |
| Bronchodilator reversibility test | 123 (34) | 63 (48) | 13 (25) | 13 (27) | 5 (14) | 29 (30) | <0.001 |
| Methacholine challenge | 53 (15) | 22 (17) | 1 (2) | 8 (17) | 6 (17) | 16 (17) | 0.107 |
| Exercise challenge test | 23 (6) | 8 (6) | 1 (2) | 2 (4) | 2 (6) | 10 (10) | 0.370* |
| FeNO | 266 (73) | 115 (87) | 25 (49) | 33 (69) | 25 (69) | 68 (71) | <0.001 |
| Nasal NO | 49 (14) | 11 (8) | 17 (34) | 8 (17) | 5 (14) | 8 (8) | <0.001 |
| Any allergy test (skin prick test or specific IgE or ImmunoCAP Rapid) | 253 (70) | 98 (74) | 30 (59) | 36 (75) | 25 (69) | 64 (67) | 0.267 |
| Chest x-ray | 92 (25) | 21 (16) | 15 (29) | 13 (27) | 13 (36) | 30 (31) | 0.030 |
| Microbiology<sup>°</sup> | 24 (7) | 4 (3) | 8 (16) | 2 (4) | 0 (0) | 10 (10) | 0.006* |
| Sweat test | 19 (5) | 4 (3) | 4 (8) | 0 (0) | 3 (8) | 8 (8) | 0.076* |

Diagnostic investigations include tests performed at any visit, i.e. at the first clinical visit or follow-up visits. FeNO: fractional exhaled nitric oxide; Nasal NO: nasal nitric oxide; p-values are calculated using chi-squared test; \*p-value calculated using Fishers exact test;
<sup>°</sup>microbiology includes throat swabs, nasal swabs, sputum, broncho-alveolar lavage

**Table 3:** Diagnostic investigations performed overall, and stratified by age.

| Diagnostic tests done | Total | Age < 5 years | Age ≥ 5 years |  |
| --- | --- | --- | --- | --- |
|  | (N=363) | (N=131) | (N=232) |  |
|  | N (%) | N (%) | N (%) | p-value |
| Lung function (spirometry or body plethysmography) | 257 (71) | 32 (24) | 225 (97) | <0.001 |
| Bronchodilator reversibility test | 123 (34) | 10 (8) | 113 (49) | <0.001 |
| Methacholine challenge | 53 (15) | 0 (0) | 53 (23) | <0.001 |
| Exercise challenge test | 23 (6) | 1 (1) | 22 (9) | 0.001 |
| FeNO | 266 (73) | 49 (37) | 217 (94) | <0.001 |
| Nasal NO | 49 (14) | 20 (15) | 29 (13) | 0.459 |
| Any allergy tests (skin prick test or specific IgE or ImmunoCAP rapid) | 253 (70) | 76 (58) | 177 (76) | <0.001 |
| Chest x-ray | 92 (25) | 56 (43) | 36 (16) | <0.001 |
| Microbiology<sup>°</sup> | 24 (7) | 11 (8) | 13 (6) | 0.304 |
| Sweat test | 19 (5) | 8 (6) | 11 (5) | 0.575 |

Diagnostic investigations include tests performed at any visit, i.e. at the first clinical visit or follow-up visits. FeNO: fractional exhaled nitric oxide; Nasal NO: nasal nitric oxide; <sup>°</sup>microbiology includes throat swabs, nasal swabs, sputum, broncho-alveolar lavage; p-values are calculated using chi-squared test

### 3.4 Treatment

Almost 90% (321 of the 363 children in the study) were prescribed treatment, which varied by diagnosis (Table 4). Inhaled asthma medication including bronchodilators and ICS was the treatment most often prescribed, for 216 of all children (60%). Inhaled corticosteroids alone or in combination with LABA were mostly prescribed for 111 of the 132 children diagnosed with asthma and asthma-like conditions (84%). Antibiotics were prescribed for about one-tenth of all children, most commonly for 43% of the children with RTI, and especially for those diagnosed with PBB, 81% of whom received antibiotics. Among children diagnosed with UACS, 83% were prescribed nasal corticosteroids. Oral corticosteroids and antireflux medications were rarely used (2% and 3% respectively). No antitussives were prescribed except for one child for whom a trial of codeine-containing antitussive was recommended in case of progressive cough episodes.

**Table 4:** Treatment prescribed in the entire study population and stratified by diagnostic group.

| Treatment prescribed | Total | Asthma and<br>asthma-like<br>conditions | Recurrent /<br>protracted<br>infections | Upper airway<br>cough<br>syndrome | Post-<br>infectious<br>cough | Unknown<br>etiology or<br>other |  |
| --- | --- | --- | --- | --- | --- | --- | --- |
|  | (N=363) | (N=132) | (N=51) | (N=48) | (N=36) | (N=96) |  |
|  | N (%) | N (%) | N (%) | N (%) | N (%) | N (%) | p-value |
| Any treatment | 321 (88) | 132 (100) | 43 (84) | 43 (90) | 26 (72) | 77 (80) | <0.001 |
| Asthma inhaled<br>medication<br>(bronchodilator or any<br>ICS) | 216 (60) | 122 (92) | 19 (37) | 8 (17) | 18 (50) | 49 (51) | <0.001 |
| Bronchodilator | 159 (44) | 92 (70) | 18 (35) | 8 (17) | 13 (36) | 28 (29) | <0.001 |
| ICS | 120 (33) | 71 (54) | 6 (12) | 5 (10) | 14 (39) | 24 (25) | <0.001 |
| ICS + LABA | 65 (18) | 47 (36) | 2 (4) | 0 (0) | 2 (6) | 14 (15) | <0.001 |
| Any ICS | 175 (48) | 111 (84) | 8 (16) | 4 (8) | 16 (44) | 36 (38) | <0.001 |
| LTRA | 32 (9) | 12 (9) | 1 (2) | 5 (10) | 4 (11) | 10 (10) | 0.361* |
| Oral corticosteroid | 7 (2) | 6 (5) | 0 (0) | 0 (0) | 1 (3) | 0 (0) | 0.076* |
| Antihistamine | 29 (8) | 22 (17) | 0 (0) | 2 (4) | 0 (0) | 5 (5) | <0.001* |
| Antibiotic | 40 (11) | 3 (2) | 22 (43) | 3 (6) | 1 (3) | 11 (11) | <0.001 |
| Antireflux medication | 12 (3) | 3 (2) | 2 (4) | 0 (0) | 0 (0) | 7 (7) | 0.122* |
| Nasal corticosteroid | 159 (44) | 57 (43) | 13 (25) | 40 (83) | 9 (25) | 40 (42) | <0.001 |

Treatment prescribed include treatment prescribed at any visit, i.e. at the first clinical visit or follow-up visits. ICS: inhaled corticosteroids; LABA: long acting beta agonist; LTRA: leukotriene receptor antagonist; p-values calculated using chi-square test; \*p-value calculated using Fishers exact test

## 4. DISCUSSION

We examined and reported the diagnostic investigations, the final diagnosis, and treatment prescribed by pediatric pulmonologists for children visiting respiratory outpatient clinics in Switzerland for prolonged or recurrent cough. These clinicians made a wide range of diagnoses, with asthma being most common. Diagnosis differed strongly by age; asthma was diagnosed 3.5 times more often in schoolchildren while RTI, including PBB, was diagnosed 3 times more often in preschoolers.

### 4.1 Strengths and limitations

This is the first study of diagnostic evaluation of children with prolonged or recurrent cough in clinical settings in Switzerland; it is among the first in Europe and one of the largest of such studies to capture the spectrum of diagnoses. The multicenter design included centers across Switzerland which makes the results more likely to be generalizable. Additionally, our study’s broad inclusion criterion—children newly referred for any prolonged or recurrent cough—allowed us to study a wide spectrum of children with troublesome cough referred to a specialist. This study’s limitations include the fact that since the SPAC study is observational, diagnostic evaluations were not standardized according to a predefined diagnostic algorithm for cough, and final diagnosis was not based on diagnostic criteria defined a priori. Nevertheless, all participating pediatric pulmonologists are board-certified and based their diagnoses on a combination of history, clinical signs, and objective diagnostic tests. Finally, though our study was based in centers across Switzerland, our study population included families who spoke German or French; its results might therefore not be fully generalizable to other demographic groups that, for example, include recent immigrants.

### 4.2 Comparison with other studies

The diagnostic spectrum reported for children presenting with chronic cough in other clinical studies is summarized in Table S4 [1, 3, 15-27]. Differences between studies may be due to variation in study period, geographical location, referral practices, clinical settings (general vs more specialized centers), number of centers (single vs multicenter), inclusion criteria such as age range and definition of chronic cough (ranging from more than 3 weeks to more than 8 weeks), exclusion criteria (e.g., history of respiratory tract infections in preceding weeks), diagnostic evaluation (use of predefined cough diagnostic algorithm [7, 33-36], standard diagnostic criteria and standard investigations vs retrospective review of medical records), and follow-up period.

Asthma and asthma-like conditions were often the most frequent diagnoses, given to 28 to 59% of children seen for cough [3, 15, 16, 19-24]. We found the same (36%). Contrary to our expectations, PBB was diagnosed rather rarely (7%). In other studies, this diagnosis was given to 10 to 41% of the children [1, 18, 19, 21-23, 26]. In two Australian studies by Marchant et al.[18] and Chang et al.[1], PBB was the most frequent diagnosis (≥ 40%) overall. However, their study population was younger (average age < 5 years)—PBB is known mainly as a disease of preschoolers—and included children with aboriginal descent, in whom PBB is more common [37].

### 4.3 Interpretation of findings and future studies

Asthma, including asthma-like conditions, was the most common diagnosis in our study, given to 36% overall and to 42% of school-age children. Overall, 60% of the children were prescribed inhaled asthma treatment (bronchodilators and/or ICS) and 48% were prescribed any ICS, which is higher than the proportion of children diagnosed with asthma or asthma-like conditions (36%). According to the European Respiratory Society (ERS) guidelines for the diagnosis of asthma in children aged 5 to 16 years, asthma treatment should only be given in case of a confirmed asthma diagnosis or in case of a treatment trial when asthma is suspected [38]. Hence, the higher proportion of prescribed asthma treatment in this study might reflect treatment trials in unconfirmed cases where asthma was suspected as an alternative diagnosis. Of clinical interest is the fact that parents reported wheeze in only 62% of children later diagnosed with asthma or asthma-like conditions, which implies that suspicion of asthma should remain elevated among children seen for prolonged or recurrent cough, even in the absence of parent-reported wheeze, and that objective tests should be used to support an asthma diagnosis.

In addition to asthma, a wide range of other diagnoses must be considered in children with prolonged or recurrent cough. Unlike in adults, GERD was rarely diagnosed (2%) in both preschoolers and schoolchildren. This may be due in part to the absence of a standard diagnostic test for the diagnosis of GERD in children [39]. In a fifth of the children, the diagnosis remained unclear, highlighting the diagnostic challenge but probably also the absence of consensus on how to label some diagnoses such as nonspecific chronic cough with natural resolution. A randomized controlled trial has shown that a standardized diagnostic algorithm improves clinical outcomes in children presenting with cough and helps to determine the primary diagnosis in 99.6% of the cases [40]. It is thus worthwhile to standardize diagnostic investigation and the terminology used for cough diagnoses in Switzerland. This process has now started in Switzerland as part of a project funded by the Swiss Personalized Health Network (SPHN) initiative—the SPHN-SPAC project.

## 5. CONCLUSION

The wide spectrum of diagnoses given to children presenting with prolonged or recurrent cough in Switzerland emphasizes the importance of comprehensive consideration and investigation of diagnoses beyond asthma in children with cough. Although asthma was the most frequent diagnosis observed in the study, it was reached in only 39% of school-age children, and a significant difference was observed between school-age children and preschoolers, among whom asthma was diagnosed in 11%. Because no definitive diagnosis could be made in 20% of children evaluated for prolonged or recurrent cough, standardizing both diagnostic investigation and labelling of cough diagnoses, which may help improve the diagnostic yield in Switzerland, should be explored.

## AUTHOR CONTRIBUTIONS

MCM, AE, ESLP and CK conceptualize this analysis. CdJ, BK, AM, NR, JU, OS, and CK recruited participants for the study. MCM, AE, SG, TK, CdJ, and ESPL collected and entered data. MCM analyzed the data. MCM drafted the manuscript. All authors interpreted the data, critically reviewed the manuscript, and approved the final version of the manuscript.

## ACKNOWLEDGEMENTS

The authors would like to thank Dr Christopher Ritter, PhD (Institute for Social and Preventive Medicine, University of Bern) for his editorial contribution, the families who took part in the SPAC study, the research assistants (Natalie Messerli, Gia Thu Ly, Labinata Gjokaj and Meret Ryser) for helping with the data collection and data entry, PedNet Bern for supporting data collection in Bern, and the members of the SPAC Study Team.

## THE SPAC STUDY TEAM

Members of the SPAC Study Team are: D. Mueller-Suter, P. Eng, and B. Kern (Canton Hospital Aarau, Aarau, Switzerland); U. Frey, J. Hammer, A. Jochmann, D. Trachsel, J. Usemann and A. Oettlin (University Children’s Hospital Basel, Basel, Switzerland); P. Latzin, C. Casaulta, C. Abbas, M. Bullo, O. Fuchs, E. Kieninger, I. Korten, L. Krüger, B. Seyfried, S. Yammine, and C.C.M de Jong (University Children’s Hospital Bern, Bern, Switzerland); P. Iseli (Children’s Hospital Chur, Chur, Switzerland); K. Hoyler (private pediatric pulmonologist, Horgen, Switzerland); S. Blanchon, S. Guerin, and I. Rochat (University Children’s Hospital Lausanne, Lausanne, Switzerland); N. Regamey, M. Lurà, M. Hitzler, K. Hrup, and J. Stritt (Canton Hospital Lucerne, Lucerne, Switzerland); J. Barben (Children’s Hospital St Gallen, St Gallen, Switzerland); O. Sutter (private pediatric practice, Worb, Bern, Switzerland); A. Moeller, A. Hector, K. Heschl, A. Jung, T. Schürmann, and L. Thanikkel (University Children’s Hospital Zurich, Zurich, Switzerland); and C.E. Kuehni, C. Ardura-Garcia, D. Berger, S. Glick, B. Guerra, T. Krasnova, R. Makhoul, M.C. Mallet, E. Pedersen, and M. Goutaki (Institute of Social and Preventive Medicine, University of Bern).

## DATA AVAILABILITY STATEMENT

The data that support the findings of this study are available on reasonable request from the corresponding author.

## REFERENCES

1. Chang AB, Robertson CF, Van Asperen PP, Glasgow NJ, Mellis CM, Masters IB, Teoh L, Tjhung I, Morris PS, Petsky HL, Willis C, Landau LI. A Multicenter Study on Chronic Cough in Children: Burden and Etiologies Based on a Standardized Management Pathway. Chest. 2012;142(4):943–50.

2. Marchant JM, Newcombe PA, Juniper EF, Sheffield JK, Stathis SL, Chang AB. What is the burden of chronic cough for families? Chest. 2008;134(2):303–9.

3. Callahan CW. Etiology of chronic cough in a population of children referred to a pediatric pulmonologist. J Am Board Fam Pract. 1996;9(5):324–7.

4. Morice AH, Millqvist E, Bieksiene K, Birring SS, Dicpinigaitis P, Domingo Ribas C, Hilton Boon M, Kantar A, Lai K, McGarvey L, Rigau D, Satia I, Smith J, Song WJ, Tonia T, van den Berg JWK, van Manen MJG, Zacharasiewicz A. ERS guidelines on the diagnosis and treatment of chronic cough in adults and children. The European respiratory journal. 2020;55(1).

5. Weinberger M, Fischer A. Differential diagnosis of chronic cough in children. Allergy Asthma Proc. 2014;35(2):95–103.

6. Haydour Q, Alahdab F, Farah M, Barrionuevo P, Vertigan AE, Newcombe PA, Pringsheim T, Chang AB, Rubin BK, McGarvey L, Weir KA, Altman KW, Feinstein A, Murad MH, Irwin RS. Management and diagnosis of psychogenic cough, habit cough, and tic cough: a systematic review. Chest. 2014;146(2):355–72.

7. Chang AB, Glomb WB. Guidelines for evaluating chronic cough in pediatrics: ACCP evidence-based clinical practice guidelines. Chest. 2006;129(1 Suppl):260s-83s.

8. Waring G, Kirk S, Fallon D. The impact of chronic non-specific cough on children and their families: A narrative literature review. J Child Health Care. 2020;24(1):143–60.

9. Chang AB, Oppenheimer JJ, Weinberger M, Grant CC, Rubin BK, Irwin RS. Etiologies of Chronic Cough in Pediatric Cohorts: CHEST Guideline and Expert Panel Report. Chest. 2017;152(3):607–17.

10. Faniran AO, Peat JK, Woolcock AJ. Persistent cough: is it asthma? Arch Dis Child. 1998;79(5):411–4.

11. Divaret-Chauveau A, Mauny F, Hose A, Depner M, Dalphin ML, Kaulek V, Barnig C, Schaub B, Schmausser-Hechfellner E, Renz H, Riedler J, Pekkanen J, Karvonen AM, Täubel M, Lauener R, Roduit C, Vuitton DA, von Mutius E, Demoulin-Alexikova S. Trajectories of cough without a cold in early childhood and associations with atopic diseases. Clin Exp Allergy. 2022.

12. Jurca M, Goutaki M, Latzin P, Gaillard EA, Spycher BD, Kuehni CE. Isolated night cough in children: how does it differ from wheeze? ERJ Open Res. 2020;6(4).

13. Kantar A, Seminara M. Why chronic cough in children is different. Pulm Pharmacol Ther. 2019;56:51–5.

14. Kantar A, Chang AB, Shields MD, Marchant JM, Grimwood K, Grigg J, Priftis KN, Cutrera R, Midulla F, Brand PLP, Everard ML. ERS statement on protracted bacterial bronchitis in children. The European respiratory journal. 2017;50(2).

15. Holinger LD. Chronic cough in infants and children. Laryngoscope. 1986;96(3):316–22.

16. Khoshoo V, Edell D, Mohnot S, Haydel R, Jr., Saturno E, Kobernick A. Associated factors in children with chronic cough. Chest. 2009;136(3):811–5.

17. Cash H, Trosman S, Abelson T, Yellon R, Anne S. Chronic cough in children. JAMA Otolaryngol Head Neck Surg. 2015;141(5):417–23.

18. Marchant JM, Masters IB, Taylor SM, Cox NC, Seymour GJ, Chang AB. Evaluation and outcome of young children with chronic cough. Chest. 2006;129(5):1132–41.

19. Asilsoy S, Bayram E, Agin H, Apa H, Can D, Gulle S, Altinoz S. Evaluation of chronic cough in children. Chest. 2008;134(6):1122–8.

20. Karabel M, Kelekçi S, Karabel D, Gürkan MF. The evaluation of children with prolonged cough accompanied by American College of Chest Physicians guidelines. Clin Respir J. 2014;8(2):152–9.

21. Usta Guc B, Asilsoy S, Durmaz C. The assessment and management of chronic cough in children according to the British Thoracic Society guidelines: descriptive, prospective, clinical trial. Clin Respir J. 2014;8(3):330–7.

22. Gedik AH, Cakir E, Torun E, Demir AD, Kucukkoc M, Erenberk U, Uzuner S, Nursoy M, Ozkaya E, Aksoy F, Gokce S, Bahali K. Evaluation of 563 children with chronic cough accompanied by a new clinical algorithm. Italian Journal of Pediatrics. 2015;41(1):73.

23. Ilarslan NEC, Gunay F, Haskologlu ZS, Bal SK, Tezcaner ZC, Kirsaclioglu CT, Firat S, Altuntas C, Ciftci B, Ozgursoy OB, Cobanoglu N. Evaluation of children with chronic cough including obstructive sleep apnea: a single-center experience. Eur J Pediatr. 2019;178(2):189–97.

24. Yilmaz Topal O. Evaluation of Chronic Cough Etiologies in Children. Turkish Journal of Pediatric Disease. 2023.

25. Chen X, Peng WS, Wang L. Etiology analysis of nonspecific chronic cough in children of 5 years and younger. Medicine (Baltimore). 2019;98(3):e13910.

26. Yu X, Kong L, Jiang W, Dai Y, Wang Y, Huang L, Luo W, Lai K, Hao C. Etiologies associated with chronic cough and its clinical characteristics in school-age children. Journal of thoracic disease. 2019;11(7):3093–102.

27. Thomson F, Masters IB, Chang AB. Persistent cough in children and the overuse of medications. J Paediatr Child Health. 2002;38(6):578–81.

28. Pedersen ESL, de Jong CCM, Ardura-Garcia C, Barben J, Casaulta C, Frey U, Jochmann A, Latzin P, Moeller A, Regamey N, Singer F, Spycher B, Sutter O, Goutaki M, Kuehni CE. The Swiss Paediatric Airway Cohort (SPAC). ERJ Open Res. 2018;4(4).

29. Chang AB, Landau LI, Van Asperen PP, Glasgow NJ, Robertson CF, Marchant JM, Mellis CM. Cough in children: definitions and clinical evaluation. Med J Aust. 2006;184(8):398–403.

30. ATS/ERS recommendations for standardized procedures for the online and offline measurement of exhaled lower respiratory nitric oxide and nasal nitric oxide, 2005. Am J Respir Crit Care Med. 2005;171(8):912-30.

31. Crapo RO, Casaburi R, Coates AL, Enright PL, Hankinson JL, Irvin CG, MacIntyre NR, McKay RT, Wanger JS, Anderson SD, Cockcroft DW, Fish JE, Sterk PJ. Guidelines for methacholine and exercise challenge testing-1999. This official statement of the American Thoracic Society was adopted by the ATS Board of Directors, July 1999. Am J Respir Crit Care Med. 2000;161(1):309-29.

32. Miller MR, Hankinson J, Brusasco V, Burgos F, Casaburi R, Coates A, Crapo R, Enright P, van der Grinten CP, Gustafsson P, Jensen R, Johnson DC, MacIntyre N, McKay R, Navajas D, Pedersen OF, Pellegrino R, Viegi G, Wanger J. Standardisation of spirometry. The European respiratory journal. 2005;26(2):319–38.

33. Irwin RS, Curley FJ, French CL. Chronic cough. The spectrum and frequency of causes, key components of the diagnostic evaluation, and outcome of specific therapy. Am Rev Respir Dis. 1990;141(3):640–7.

34. Irwin RS, Boulet LP, Cloutier MM, Fuller R, Gold PM, Hoffstein V, Ing AJ, McCool FD, O’Byrne P, Poe RH, Prakash UB, Pratter MR, Rubin BK. Managing cough as a defense mechanism and as a symptom. A consensus panel report of the American College of Chest Physicians. Chest. 1998;114(2 Suppl Managing):133s-81s.

35. Chang AB, Robertson CF, van Asperen PP, Glasgow NJ, Masters IB, Mellis CM, Landau LI, Teoh L, Morris PS. Can a management pathway for chronic cough in children improve clinical outcomes: protocol for a multicentre evaluation. Trials. 2010;11:103.

36. Shields MD, Bush A, Everard ML, McKenzie S, Primhak R. BTS guidelines: Recommendations for the assessment and management of cough in children. Thorax. 2008;63 Suppl 3:iii1-iii15.

37. Laird P, Totterdell J, Walker R, Chang AB, Schultz A. Prevalence of chronic wet cough and protracted bacterial bronchitis in Aboriginal children. ERJ Open Res. 2019;5(4).

38. Gaillard EA, Kuehni CE, Turner S, Goutaki M, Holden KA, de Jong CCM, Lex C, Lo DKH, Lucas JS, Midulla F, Mozun R, Piacentini G, Rigau D, Rottier B, Thomas M, Tonia T, Usemann J, Yilmaz O, Zacharasiewicz A, Moeller A. European Respiratory Society clinical practice guidelines for the diagnosis of asthma in children aged 5-16 years. The European respiratory journal. 2021;58(5).

39. Rosen R, Vandenplas Y, Singendonk M, Cabana M, DiLorenzo C, Gottrand F, Gupta S, Langendam M, Staiano A, Thapar N, Tipnis N, Tabbers M. Pediatric Gastroesophageal Reflux Clinical Practice Guidelines: Joint Recommendations of the North American Society for Pediatric Gastroenterology, Hepatology, and Nutrition and the European Society for Pediatric Gastroenterology, Hepatology, and Nutrition. J Pediatr Gastroenterol Nutr. 2018;66(3):516–54.

40. Chang AB, Robertson CF, van Asperen PP, Glasgow NJ, Masters IB, Teoh L, Mellis CM, Landau LI, Marchant JM, Morris PS. A cough algorithm for chronic cough in children: a multicenter, randomized controlled study. Pediatrics. 2013;131(5):e1576–83.

