## Supplementary Tables and Figures for "Diagnosis in children with prolonged or recurrent cough: findings from the Swiss Paediatric Airway Cohort"

**Title:**

Maria Christina Mallet^1,2^, Annina Elmiger^1^, Sarah Glick^1^, Tayisiya Krasnova^1^, Carmen CM de Jong^3^, Barbara Kern^4^, Alexander Moeller^5^, Nicolas Regamey^6^, Oliver Sutter^7^, Jakob Usemann^8^, SPAC Study Team^9^, Eva SL Pedersen^1^, Claudia E Kuehni^1,3^

**Affiliations**

1. Institute of Social and Preventive Medicine, University of Bern, Bern, Switzerland
2. Graduate School for Health Sciences, University of Bern, Bern, Switzerland
3. Division of Paediatric Respiratory Medicine, Department of Paediatrics, Inselspital, Bern University Hospital, University of Bern, Bern, Switzerland
4. Department of Paediatrics, Kantonsspital Aarau, Aarau, Switzerland
5. Department of Respiratory Medicine, University Children’s Hospital Zurich and Children’s Research Center, University of Zurich, Zurich, Switzerland
6. Division of Paediatric Pulmonology, Children's Hospital, Cantonal Hospital Lucerne, Lucerne, Switzerland
7. Paediatric Practice, Worb, Switzerland
8. University Children’s Hospital Basel UKBB, Basel, Switzerland
9. A list of the SPAC Study Team can be found in the acknowledgements section

**Corresponding author**: Prof. Claudia E. Kuehni, Institute of Social and Preventive Medicine, University of Bern, Mittelstrasse 43, 3012, Bern, Switzerland; Tel: +41 31 684 35 07;

**Supplementary Tables and Figures**

**Table S1:** Description of variables and number and percentage of missing values

| **Variable** | **Question** | **Missing, N (%)** |
| --- | --- | --- |
| **Sociodemographic** |  |  |
| Sex |  | 0 |
| Age |  | 0 |
| Country of origin | In which country was the child born? | 0 |
| Maternal highest educational level | What training did the mother of the child complete? | 10 (3) |
| Paternal highest educational level | What training did the father of the child complete? | 13 (4) |
| **Environmental exposure** |  |  |
| Parental smoking | Does the mother/father smoke? | 12 (3) |
| **Parental history** |  |  |
| Asthma | Did the child’s mother or father ever have asthma or need an asthma spray to inhale? | 0 |
| Hay fever | Did the child’s mother or father ever have hay fever? | 0 |
| Chronic cough | Did the child’s mother or father ever have chronic cough or bronchitis? | 0 |
| **Cough symptoms and characteristics in the past 12 months** |  |  |
| Cough with colds | Does your child usually have a cough with colds? | 3 (1)  Missing recoded to No |
| Cough without a cold | Does your child have a cough even without having a cold? | 5 (1)  Missing recoded to No |
| Dry night cough | In the last 12 months has your child had a dry cough at night, apart from a cough associated with a cold or a chest infection? | 6 (2)  Missing recoded to No |
| Cough more than others | Do you think your child coughs more than other children? | 5 (1)  Missing recoded to No |
| Cough > 4 weeks | Has your child had a cough in the past 12 months that lasted more than 4 weeks in a row? | 2 (1)  Missing recoded to No |
| Cough > 2 months | Has your child had a cough in the past 12 months that lasted more than 2 months in a row? | 20 (6)  Missing recoded to No |
| Cough type | Most of the time, is your child’s cough rather dry or wet? | 1 (0) |
| Cough triggers | Which of the following situation has caused your child to experience coughing in the last 12 months? |  |
|  | Cold | 33 (9)  Missing recoded to No |
|  | Exercise/play | 32 (9)  Missing recoded to No |
|  | Pollen | 86 (24)  Missing recoded to No |
|  | Pets | 80 (22)  Missing recoded to No |
|  | House dust | 90 (25)  Missing recoded to No |
|  | Food/drinks | 77 (21)  Missing recoded to No |
|  | Cold air | 69 (19)  Missing recoded to No |
|  | Change in temperature/weather | 82 (23)  Missing recoded to No |
|  | Laughter/crying | 58 (16)  Missing recoded to No |
| **Wheeze & other symptoms** |  |  |
| Wheeze in past 12 months | Did your child have wheezing or whistling in the chest in the past 12 months? | 0 |
| Number of colds ≥ 7/year | How many colds did your child have in the past 12 months? | 6 (2)  Missing recoded with median category |
| Otitis media | Did your child have a middle ear infection in the past 12 months? | 6 (2)  Missing recoded to No |
| Snoring (without cold/almost always) | During the past 12 months, has your child sometimes snored? | 3 (1)  Missing recoded to No |
| Pneumonia | Did your child have a pneumonia in the last 12 months? | 4 (1)  Missing recoded to No |
| Rhino-conjunctivitis | In the past 12 months, has your child had a problem with sneezing, or a runny, or blocked nose when he/she did not have a cold or the flu? If yes, in the past 12 months, has this nose problem been accompanied by itchy-watery eyes? | 21 (6)  Missing recoded to No |

**Figure S1:** Flowchart of study participants

2206 children with signed informed consent, completed baseline parental questionnaire, and at least one outpatient clinical letter by September 2022

1843 children excluded:

- 1754 where prolonged or recurrent cough was not the main reason for visit at the outpatient clinic
- 83 were seen in follow-up for prolonged or recurrent cough (not new referrals)
- 6 were already known asthma prior to referral

363 children seen for prolonged or recurrent cough as main reason and included in the analysis

**Table S2:** Final diagnosis given by pediatric pulmonologists for children seen for prolonged or recurrent cough in the study population (N=363) overall and stratified by age

| **Diagnosis** | **Total** | **< 5 years** | **≥ 5 years** |
| --- | --- | --- | --- |
|  | **(N=363)** | **(N=131)** | **(N=232)** |
|  | **N (%)** | **N (%)** | **N (%)** |
| Asthma | 104 (29) | 14 (11) | 90 (39) |
| Asthma-like conditions* | 28 (8) | 20 (15) | 8 (3) |
| Recurrent respiratory tract infections | 25 (7) | 16 (12) | 9 (4) |
| Protracted bacterial bronchitis | 26 (7) | 17 (13) | 9 (4) |
| Upper airway cough syndrome/chronic obstructive rhinopathy with posterior rhinorrhea/postnasal drip | 48 (13) | 19 (15) | 29 (13) |
| Postinfectious cough with hyperactivity of cough receptors/bronchial hyperreactivity | 36 (10) | 14 (11) | 22 (9) |
| Somatic/tic cough disorder | 6 (2) | 0 (0) | 6 (3) |
| Gastroesophageal reflux | 2 (1) | 1 (1) | 1 (0) |
| Other ^#^ | 15 (4) | 2 (2) | 13 (6) |
| Unknown/unclear etiology | 73 (20) | 28 (21) | 45 (19) |

*Asthma-like conditions include cough variant asthma, episodic viral wheeze, and recurrent obstructive bronchitis

^#^Other includes house dust mite allergy and other atopic cough (n=8), exercise induced laryngeal obstruction (n=1), functional respiratory disorder (n=1), physical deconditioning (n=1), tracheomalacia (n=1), obstructive sleep apnea syndrome (n=1), adenoid hyperplasia (n=1), and non-cystic fibrosis bronchiectasis (n=1)

**Table S3:** Parent-reported characteristics and symptoms in the entire study population, and stratified by diagnostic group (sensitivity analysis with complete case analysis)

|  | **Total**  **(N=363)** | **Asthma and asthma-like conditions**  **(N=132)** | **Recurrent/**  **protracted infection**  **(N=51)** | **Upper airway cough syndrome**  **(N=48)** | **Postinfectious cough**  **(N=36)** | **Unknown etiology or other**  **(N=96)** | **p-value** |
| --- | --- | --- | --- | --- | --- | --- | --- |
|  | **N (%)** | **N (%)** | **N (%)** | **N (%)** | **N (%)** | **N (%)** |  |
| **Cough symptoms and characteristics in the past 12 months** |  |  |  |  |  |  |  |
| Cough with colds (N=360) | 355 (99) | 128 (98) | 50 (100) | 46 (98) | 35 (97) | 96 (100) | 0.335^#^ |
| Cough without a cold (N=358) | 319 (89) | 114 (88) | 39 (78) | 45 (96) | 31 (89) | 90 (94) | 0.028 |
| Dry night cough (N=357) | 262 (73) | 92 (71) | 29 (58) | 37 (79) | 29 (81) | 75 (79) | 0.050 |
| Cough more than others (N=358) | 328 (92) | 119 (90) | 45 (88) | 47 (98) | 29 (88) | 88 (93) | 0.272^#^ |
| Cough > 4 weeks (N=361) | 282 (78) | 99 (75) | 35 (70) | 40 (83) | 30 (83) | 78 (82) | 0.298 |
| Cough > 2 months (N=343) | 178 (52) | 42 (34) | 26 (54) | 29 (63) | 23 (66) | 58 (63) | <0.001 |
| Cough type (N=362) |  |  |  |  |  |  |  |
| Mostly dry | 157 (43) | 54 (41) | 11 (22) | 26 (54) | 15 (42) | 51 (53) | <0.001 |
| Mostly wet | 44 (12) | 11 (8) | 18 (35) | 4 (8) | 3 (8) | 8 (8) |  |
| Both dry and wet | 161 (44) | 66 (50) | 22 (43) | 18 (38) | 18 (50) | 37 (39) |  |
| Cough triggers |  |  |  |  |  |  |  |
| Any aeroallergen | 126 (35) | 63 (48) | 11 (22) | 15 (31) | 6 (17) | 31 (32) | 0.001 |
| Any physical trigger (exercise/laughter/crying) | 273 (75) | 101 (77) | 37 (73) | 36 (75) | 26 (72) | 73 (76) | 0.971 |
| Cold air/ temperature change | 196 (54) | 74 (56) | 25 (49) | 28 (58) | 23 (64) | 46 (48) | 0.413 |
| **Wheeze & other symptoms in past 12 months** |  |  |  |  |  |  |  |
| Wheeze | 172 (47) | 82 (62) | 24 (47) | 20 (42) | 12 (33) | 34 (35) | <0.001 |
| Number of colds ≥7/year (N=357) | 110 (31) | 30 (23) | 26 (53) | 17 (37) | 10 (28) | 27 (28) | 0.002 |
| Otitis media (N=357) | 64 (18) | 21 (16) | 13 (25) | 11 (23) | 4 (12) | 15 (16) | 0.379 |
| Snoring° (N=360) | 144 (40) | 56 (42) | 12 (24) | 25 (52) | 13 (37) | 38 (40) | 0.069 |
| Pneumonia (N=359) | 44 (12) | 18 (14) | 6 (12) | 3 (6) | 4 (11) | 13 (14) | 0.701 |
| Rhino-conjunctivitis (N=337) | 77 (23) | 41 (33) | 2 (5) | 8 (19) | 4 (12) | 22 (24) | 0.001 |

°without cold/almost always; *p-value calculated with kwallis test; p-value calculated with ^#^Fishers exact test

**Table S4:** Comparison of setting, inclusion criteria, diagnostic evaluation and frequency of diagnosis in studies investigating etiology of chronic cough in clinical studies of children

| **Study** | **Country** | **Setting and study population (inclusion and exclusion criteria)** | **Methods/diagnostic evaluation** | **Frequency of diagnosis** |
| --- | --- | --- | --- | --- |
| Holinger, Laryngoscope, 1986 [1] | United States | *Inclusion:* Children aged less than 16 years having chronic cough (cough >4 weeks) as chief complaint, with a normal chest X-ray, having been evaluated and treated by at least one referring physician, and seen between January 1980 and June 1984 in the department of Otolaryngology-Head-and Neck Surgery in a Children’s hospital, and in most cases were primarily referred for bronchoscopy (single center)  *Sample size*: 38 | Evaluation was individualized based on history, patient age, and based on investigations already completed prior to visit | Cough variant asthma(CVA): 39%  Sinusitis: 16%  Aberrant innominate artery: 13%  Psychogenic cough: 13%  Unknown: 13%  Subglottic stenosis: 11%  Tracheomalacia: 3%  Gastroesophageal reflux: 3%  Bronchial foreign body: 3%  Bronchogenic cyst: 3%  Most common causes by age group:  0-18 months: aberrant innominate artery and CVA  1.5-6 years: sinusitis and CVA  6-16 years: CVA and psychogenic cough |
| Callahan, J Am Board Fam Pract, 1996 [2] | Hawaii (United States) | *Inclusion:* Children referred for chronic cough (cough > 3 weeks) to the outpatient pediatric pulmonology department in a hospital between July 1993 to June 1995 (single center)  *Sample size*: 95  *Median age*: not known/specified | Retrospective review of patients’ records | Asthma: 59%  Chronic sinusitis: 9%  Gastroesophageal reflux: 9%  Chronic bronchitis: 6%  Allergic rhinitis:4%  Postbronchiolitic airway reactivity: 4%  Tracheomalacia: 3%  Psychogenic cough: 2%  Vascular ring: 1%  Hypersensitivity pneumonitis: 1% |
| Thomson, J. Paediatr Child Health, 2002 [3] | Australia | *Inclusion:* Newly referred children (4 months to 14 years) seen for persistent cough (≥ 4 weeks) by respiratory physicians at a tertiary pediatric respiratory clinic from January 1998 to January 1999 (single center)  *Sample size*: 49  *Median age*: 49 months | Retrospective chart review | Lower airway malacia disorders (tracheomalacia, bronchomalacia): 47%  Postviral cough:10%  Major airway stenosis: 10%  Postrespiratory infection cough/bronchitis: 10%  Laryngomalacia: 6%  Foreign body: 6%  Psychogenic cough: 4%  Bronchitis with tobacco smoke exposure: 4%  Post-foreign body with granulation tissue: 4%  Others (2% each): achalasia, aspiration lung disease, obstructive sleep apnea, bronchiectasis, laryngeal tracheal cleft, non-tuberculous mycobacteria and follicular bronchiolitis  Dual diagnosis was present in 27% of the children |
| Marchant et al., Chest, 2006 [4] | Australia | *Inclusion:* Any child (< 18 years) with chronic cough (cough > 3 weeks) of unknown etiology referred to a university hospital (single center) between June 2002 to June 2004  *Exclusion:* Infants born prematurely (i.e. < 37 weeks), children with known lung disease or other severe underlying disorders such as neurodevelopmental delay or cardiac abnormalities  *Sample size:* 108 children  *Median age*: 2.6 years | Study protocol based on an adult algorithm for chronic cough and modified for children[5, 6].  Children were followed up until cough resolution and up to a maximum of 12 months after study enrolment  Standard a priori definitions were used for diagnostic categories | Protracted bacterial bronchitis: 40%  Natural resolution: 20%  Other diagnosis (in number):  -Bronchiectasis: 6  -Uncertain: 5  -Asthma-like conditions: 4  -Habit cough: 1  -Eosinophilic disorders:4  -Aspiration disorders: 5  -B pertussis: 1  -M pneumoniae infection: 2  -Endobronchial tuberculosis: 1  -Gastroesophageal reflux: 3  -Upper airway cough syndrome: 3  -Bronchiolitis obliterans:1 |
| Asilsoy et al., Chest, 2008 [7] | Turkey | *Inclusion:* Children 6-14 years presenting to a research hospital for chronic cough (cough > 4 weeks) between November 2006 and May 2007 (single center)  *Exclusion:* Patients with premature birth, known lung disease, neuromotor growth deficiency, cardiac disease, growth deficiency, deformed chest and history of acute respiratory tract infection in the last 4 weeks  *Sample size*: 108  *Mean age*: 8 years | Study conducted based on the 2006 American College of Chest Physicians (ACCP) guidelines for chronic cough in children[8].  Patients were evaluation at intervals of 2-4 weeks until the cough has resolved.  Definition of diagnostic categories was used | Asthma plus asthma like symptoms 25%  Protracted bronchitis 23.4%  Upper airway cough syndrome (UACS) 20.3%  Protracted bronchitis plus asthma like symptoms 12%  UACS plus asthma like symptoms 7.4%  Gastroesophageal reflux 4.6%  Bronchiectasis 2.7%  Natural recovery 1.8%  Tuberculosis 0.9%  Congenital malformation 0.9%  Mycoplasma infection 0.9% |
| Khoshoo et al., Chest, 2009 [9] | United States | *Inclusion*: Children aged 5-12 years with a cough duration of > 8 weeks referred to a pediatric specialty center, born full term, neurodevelopmentally appropriate for age, absence of direct or indirect cigarette smoke exposure, absence of history of febrile or respiratory illness and no cardiac illness and able to do pulmonary function testing and methacholine challenge testing (single center)  *Exclusion:* Children with a known diagnosis of asthma, reactive airway disease or cystic fibrosis  Sample size: 40  *Mean age*: 7.8 years | Multispecialist and standard workup by pulmonologist, gastroenterologist, allergist, immunologist and otorhinolaryngologist. Extra tests were performed in selected cases | Gastroesophageal reflux disease: 27.5%  Allergy: 22.5%  Asthma: 7.5%  Cough variant asthma: 5%  Infection: 5%  Allergy and asthma: 15%  Allergy and reflux: 5%  Aspiration: 2.5%  All test results normal: 10%  Most children with allergy were found to have upper airway cough syndrome |
| Chang et al., Chest, 2012 [10] | Australia | *Inclusion:* Children aged < 18 years newly referred for chronic cough (cough > 4 weeks) from 5 major hospitals and 3 rural remote clinics (multicenter)  *Exclusion*: Children with a known chronic respiratory illness previously diagnosed by a respiratory physician or those with a confirmed diagnosed based on objective tests (e.g. cystic fibrosis and bronchiectasis)  *Sample size*: 346  *Mean age*: 4.5 years | Management according to standardized evidence based cough algorithm (adapted from the ACCP pediatric guideline)[11]  End point of study: primary diagnosis established, cough resolution or 12 months from time of enrolment  Standard definitions used for primary diagnoses | Protracted bacterial bronchitis (PBB) :41%  Asthma/Reactive airway disease: 15.9%  Resolved without specific diagnosis: 13.9%  Bronchiectasis: 9%  Tracheomalacia :6.1%  Habitual-psychogenic: 4.3%  Pertussis: 3.5%  Aspiration lung disease 2.3%  Upper airways : 1.4%  Mycoplasma : 1.4%  Pneumonia: 0.9%  Cystic fibrosis : 0.3%  There was also an overall significant difference for  primary diagnosis when children were grouped into  four age categories  Three most common diagnosis by age group:  0-2 years:  PBB (53%), Asthma/reactive airway disease (27%), resolved without specific diagnosis (11%)  >2 to ≤ 6 years: PBB (40%), asthma/reactive airway disease (19%), Bronchiectasis (14%)  >6 to ≤ 12 years: PBB (27%), resolved without specific diagnosis (20%), habitual-psychogenic (15%)  >12 years: PBB (29%), resolved without specific diagnosis (29%), habitual-psychogenic (21%) |
| Karabel et al., Clin Respir J, 2014 [12] | Turkey | *Inclusion*: Patients aged 7 months to 17 years presenting with complaints of chronic cough (cough > 4 weeks) to the Pediatric Pulmonology clinic (single center)  *Exclusion*: known neuromotor, cardiac or syndromic diseases, premature birth, and history of respiratory tract infection in the past 4 weeks.  *Sample size:* 270  *Mean age:* 6.5 years | Patients evaluated based on the ACCP guidelines[8]  All patients were followed up for at least 12 months  Standardized definition of causes | Most frequent causes:  Asthma: 27%  Asthma-like symptoms: 15.5%  GER: 10% |
| Usta et al., Clin Respir J, 2014 [13] | Turkey | *Inclusion*: Patient aged 5-16 years presenting with chronic cough (cough > 8 weeks) at the Department of Pediatric Allergy and Immunology of a research and training hospital between September 2009 and September 2010 (single center)  *Exclusion:* History of premature birth, neuromotor development retardation, development-growth retardation, chest wall deformity, smoking habit, clubbing, cardiac disease, any known chronic disease and/or pulmonary disease and inability to cooperate in lung function test  *Sample size*: 156  *Mean age:* 8.4 year | Cough assessed and managed according to the British Thoracic Society guidelines [14]  Patient re-evaluated at 2 to 4 week intervals and followed for 18 months until cough resolved  Standard definition for diagnosis | Postnasal drip syndrome plus asthma :19.2%  Postnasal drip syndrome: 18.6 %  Asthma: 12.2%  Protracted bacterial bronchitis: 12.2 %  Nonspecific isolated cough: 11.5%  Cough variant asthma: 9.6%  Psychogenic cough: 5.8 %  GERD: 3.2%  Postnasal drip syndrome + GERD: 1.9%  Nonspecific isolated cough + GERD: 1.9%  Asthma + GERD: 1.3%  Tuberculosis infection: 1.3%  Bronchiectasis: 0.6 %  Bronchiectasis + immune deficiency: 0.6% |
| Cash et al., JAMA Otolaryngol Head Neck Surg, 2015 [15] | United States | *Inclusion*: All patients < 18 years presenting with chronic cough (cough > 4 weeks) to two otolaryngology clinics in a tertiary care academic center from January 2009 to June 2013 (single center).  *Sample size*: 58  *Mean age*: 5.1 | Retrospective analysis of medical records | Infection (including upper respiratory tract infection, and/or Upper airway cough syndrome, sinusitis and lower respiratory tract infection): 34 %  Airway hyperreactivity including asthma/reactive airway disease :24%  Gastroesophageal reflux: 24%  Unresolved: 14%  Allergic rhinitis: 10%  Larnygomalacia: 9%  Habit: 7% |
| Gedik et al., Italian Journal of Paediatrics, 2015 [16] | Turkey | *Inclusion*: Children aged < 17 years admitted for chronic cough (cough > 4 weeks) for the first time in a large university hospital (division of pediatric pulmonology and pediatric allergy clinic) between October 2012- October 2013 (single center)  *Exclusion:* Children with previously known, chronic respiratory illness (asthma, cystic fibrosis, and bronchiectasis), neuromotor growth deficiency, cardiac or syndromic diseases, and premature birth.  *Sample size*: 563  *Mean age*: 5.4 years | Evaluation based on a cough algorithm developed by Chang et al. [11]  All participants were followed for at least 12 months  Standard definition of specific causes of cough | Atopic asthma :25%  Reactive airway disease (asthma like symptoms): 19%  PBB :12%  UACS 9.1%  Psychogenic cough:5.5%  non-cystic fibrosis bronchiectasis :5%  Bronchiolitis obliterans: 5%  Rhinosinusitis: 4.6%  CF: 3.6%  Tuberculosis: 3.4%  Pneumonia-bronchopneumonia: 3.2%  GERD: 2.7%  Others (<1%): tracheobronchomalacia, foreign body aspiration, spontaneous resolution, vascular ring, pulmonary hemosiderosis, tumor (ganglioneuroma)  Three most common causes per age group:  0-2 years: reactive airway disease :29.7%; atopic asthma: 19.5%; PBB (8.6%); CF (8.6%)  2-6 years: atopic asthma: 28.8%, reactive airway disease: 21.7%, PBB: 13.4%  >6 years: atopic asthma: 23.6%, psychogenic cough: 13.8%, PBB: 11.8% |
| Yu et al., Journal of Thoracic Disease, 2019 [17] | China | *Inclusion:* Children aged 6-14 years referred to a university/tertiary hospital for evaluation of chronic cough (cough >4 weeks) (single center)  *Exclusion:* previous known chronic respiratory illnesses such as bronchopulmonary dysplasia, immotile cilia syndrome, tuberculosis, asthma and bronchiectasis, and serious cardiac or systemic diseases  *Sample size*: 118  *Mean age*: 9.3 years | Use of step by step diagnostic algorithm according to ACCP guidelines[8].  Use of uniform diagnostic criteria | UACS: 31.4%  Cough variant asthma (CVA) :14.4%  Protracted bronchitis (PB) :10.2%  GERD: 5.9%  Eosinophilic bronchitis :1.7%  CVA + UACS: 31.4%  CVA + PB: 2.5%  Tic disorder (TD) + UACS :2.5% |
| Illarslan et al., European Journal of Pediatrics, 2019 [18] | Turkey | *Inclusion:* Children aged 1 month to 14 years admitted with chronic cough (cough > 4 weeks) to the primary care clinic of a tertiary hospital between January 2016 to January 2017 (single center)  *Exclusion:* previously diagnosed chronic respiratory illness by a pediatric respiratory/allergy physician (e.g. bronchiectasis, cystic fibrosis, asthma), congenital/acquired heart diseases, neuromuscular disease, immune deficiency, syndromes, prematurity (< 37 weeks), low birth weight (<2.5kg)  *Sample size:* 237  *Median age:* 5 years | Use of a standardized cough algorithm (based on ACCP 2006 guideline).[8]  Patients were examined every 2 weeks until a final diagnosis was made and complete cough resolution was achieved. Maximum period of follow up was 12 months.  Standard diagnostic definitions | Asthma: 34.5 %  Protracted bacterial bronchitis: 30.8 %  Pneumonia: 7.6%  Obstructive sleep apnea: 7.6%  Rhinosinusitis: 5.5%  Gastroesophageal reflux disease : 5.2%  Non-specific chronic cough: 5%  Adenotonsillary hypertrophy: 4.2%  Allergic rhinitis: 3.4%  Psychogenic cough: 1.3%  Tuberculosis: 0.4%  Interstitial lung disease: 0.4%  Primary ciliary dyskinesia: 0.4%  Three most frequent causes by age group:  0-2 years: PBB: 52.9%, Asthma: 17.6%, GERD: 8.8%  > 2– < 6 years: Asthma: 36.4%; PBB: 26.3%, Obstructive sleep apnea: 10.2%  ≥ 6 – < 14 years: Asthma: 38.8%, PBB: 28.2%, GERD: 7.1% |
| Chen et al., Medicine, 2019[19] | China | *Inclusion*: Children ≤ 5 years visiting the outpatient department of one hospital from January 2015 to August 2016 with cough as main or only clinical manifestation lasting more than 4 weeks (nonspecific chronic cough), without any chest X-ray abnormalities and with a follow-up of at least 3 months (single center)  *Exclusion:* presence of other symptoms besides cough, abnormal chest X-ray, serious systemic diseases  *Sample size*: 85  *Mean age*: 3.8 years | Retrospective analysis of medical records | UACS: 37.6%  CVA: 31.8%  Postinfectious cough (PIC): 18.8%  Gastroesophageal reflux cough: 3.5%  Allergic/atopic cough: 2.4 %  Three most common diagnosis by age group:  Infants (5 months and older):  UACS: 33.3%, CVA: 25.9%, PIC:22.2%  Pre-school (3-5 years):  UACS :39.7%, CAV: 34.5% and PIC: 17.2% |
| Yilmaz et al., Turkish Journal of Pediatric Disease, 2022 [20] | Turkey | *Inclusion:* Children aged 0-18 years admitted with the complaint of chronic cough (cough > 4 weeks) to the pediatric immunology and allergy outpatient clinic in a pediatric hospital between February to August 2022 (single center)  *Sample size*: 323  *Median age*: 7 | Children were evaluated based on ACCP guidelines for chronic cough.[8] | Asthma: 44.6%  Wheezy infant: 23.2%  Postinfectious cough: 16.7%  Postnasal drip syndrome: 13.3%  Gastroesophageal reflux: 1.2%  Foreign body aspiration: 0.6%  Psychogenic cough: 0.3%  Three most common causes by age group:  < 6 years:  wheezy infant: ~65%; postinfectious cough: ~ 22%, postnasal drip syndrome: ~ 10%  ≥6 years:  Asthma: ~65%; postnasal drip syndrome ~15%; postinfectious cough ~14% |

**References**

1.	Holinger LD. Chronic cough in infants and children. Laryngoscope. 1986;96(3):316-22.

2.	Callahan CW. Etiology of chronic cough in a population of children referred to a pediatric pulmonologist. J Am Board Fam Pract. 1996;9(5):324-7.

3.	Thomson F, Masters IB, Chang AB. Persistent cough in children and the overuse of medications. J Paediatr Child Health. 2002;38(6):578-81.

4.	Marchant JM, Masters IB, Taylor SM, Cox NC, Seymour GJ, Chang AB. Evaluation and outcome of young children with chronic cough. Chest. 2006;129(5):1132-41.

5.	Irwin RS, Curley FJ, French CL. Chronic cough. The spectrum and frequency of causes, key components of the diagnostic evaluation, and outcome of specific therapy. Am Rev Respir Dis. 1990;141(3):640-7.

6.	Irwin RS, Boulet LP, Cloutier MM, Fuller R, Gold PM, Hoffstein V, et al. Managing cough as a defense mechanism and as a symptom. A consensus panel report of the American College of Chest Physicians. Chest. 1998;114(2 Suppl Managing):133s-81s.

7.	Asilsoy S, Bayram E, Agin H, Apa H, Can D, Gulle S, et al. Evaluation of chronic cough in children. Chest. 2008;134(6):1122-8.

8.	Chang AB, Glomb WB. Guidelines for evaluating chronic cough in pediatrics: ACCP evidence-based clinical practice guidelines. Chest. 2006;129(1 Suppl):260s-83s.

9.	Khoshoo V, Edell D, Mohnot S, Haydel R, Jr., Saturno E, Kobernick A. Associated factors in children with chronic cough. Chest. 2009;136(3):811-5.

10.	Chang AB, Robertson CF, Van Asperen PP, Glasgow NJ, Mellis CM, Masters IB, et al. A Multicenter Study on Chronic Cough in Children: Burden and Etiologies Based on a Standardized Management Pathway. Chest. 2012;142(4):943-50.

11.	Chang AB, Robertson CF, van Asperen PP, Glasgow NJ, Masters IB, Mellis CM, et al. Can a management pathway for chronic cough in children improve clinical outcomes: protocol for a multicentre evaluation. Trials. 2010;11:103.

12.	Karabel M, Kelekçi S, Karabel D, Gürkan MF. The evaluation of children with prolonged cough accompanied by American College of Chest Physicians guidelines. Clin Respir J. 2014;8(2):152-9.

13.	Usta Guc B, Asilsoy S, Durmaz C. The assessment and management of chronic cough in children according to the British Thoracic Society guidelines: descriptive, prospective, clinical trial. Clin Respir J. 2014;8(3):330-7.

14.	Shields MD, Bush A, Everard ML, McKenzie S, Primhak R. BTS guidelines: Recommendations for the assessment and management of cough in children. Thorax. 2008;63 Suppl 3:iii1-iii15.

15.	Cash H, Trosman S, Abelson T, Yellon R, Anne S. Chronic cough in children. JAMA Otolaryngol Head Neck Surg. 2015;141(5):417-23.

16.	Gedik AH, Cakir E, Torun E, Demir AD, Kucukkoc M, Erenberk U, et al. Evaluation of 563 children with chronic cough accompanied by a new clinical algorithm. Italian Journal of Pediatrics. 2015;41(1):73.

17.	Yu X, Kong L, Jiang W, Dai Y, Wang Y, Huang L, et al. Etiologies associated with chronic cough and its clinical characteristics in school-age children. Journal of thoracic disease. 2019;11(7):3093-102.

18.	Ilarslan NEC, Gunay F, Haskologlu ZS, Bal SK, Tezcaner ZC, Kirsaclioglu CT, et al. Evaluation of children with chronic cough including obstructive sleep apnea: a single-center experience. Eur J Pediatr. 2019;178(2):189-97.

19.	Chen X, Peng WS, Wang L. Etiology analysis of nonspecific chronic cough in children of 5 years and younger. Medicine (Baltimore). 2019;98(3):e13910.

20.	Yilmaz Topal O. Evaluation of Chronic Cough Etiologies in Children. Turkish Journal of Pediatric Disease. 2023.
